# The primacy of meeting public university students’ essential needs during the COVID-19 pandemic: a new higher education priority

**DOI:** 10.1101/2021.11.11.21266220

**Authors:** Meredith Manze, Anna Lattanzio, Jenna Larsen, Julia Keegan, Nick Freudenberg, Heidi E. Jones

## Abstract

**Objectives:** We investigated the holistic experiences of university students during the pandemic.

**Participants:** 38 students in a public university system in New York City (NYC) purposively selected from neighborhoods highly affected by the pandemic based on level of self-reported impact

**Methods:** We conducted virtual in-depth interviews from May to August 2021 and analyzed data using thematic coding and constant comparison techniques informed by grounded theory.

**Results:** Financial and social support systems, such as governmental and school resources, were critical to addressing essential needs and allowing students to persist. For those whose essential needs were met, faculty members’ flexibility and students’ experience with online learning were central to their academic success.

**Conclusions:** Institutions of higher education should strengthen financial and social support systems to meet students’ essential needs. Academic policies to bolster online pedagogy and faculty’s flexibility can facilitate student retention and completion.

## Introduction

The COVID-19 pandemic disrupted all facets of the personal, professional, and academic lives of students pursuing higher education. Many college students lost employment, experienced anxiety and loss of concentration, and endured food and financial insecurity, threatening their ability to remain in school.^1^ For the most vulnerable students, pre-existing mental health and financial problems were exacerbated by the pandemic.^2^ However, most research has focused on important but narrow areas of the effects of COVID-19 on college students, such as difficulties with online learning or interpersonal relationships,^3,4^ without full consideration of the broader context and constellation of personal and social factors that may have an impact on academic persistence and success. This study examines how the pandemic affected students’ engagement in higher education, and in particular their capacity to meet their essential needs including food, housing, and financial security. Growing evidence suggests that challenges in meeting essential needs can undermine academic success.^1-3^

The City University of New York (CUNY) is the largest urban public university in the United States (U.S.), enrolling over a quarter of a million students each year across 25 campuses (granting associates, bachelors, masters, and doctoral degrees) in New York City. By some standard indicators, CUNY students are in socially and financially precarious situations. Almost half of students (44%) are the first in their family to pursue a college degree.^5^ In 2018, nearly a third of undergraduate students reported being worried about having enough food and over half had concerns about affording housing.^6^ New York City was considered an early epicenter of the COVID-19 pandemic in March 2020,^7^ causing significant losses to employment that disrupted many people’s access to a reliable source of income. Such circumstances further heightened students’ financial insecurities.^8^

At public universities, the potentially unstable nature of students’ lives and the larger social context in which they live should be considered when investigating and developing ways to support their student body. Few studies have conducted in-depth inquiries into the holistic lived experiences of students in higher education during the pandemic by assessing personal, social, and institutional factors that affect academic achievement. By better understanding student experiences one year into the pandemic, leaders of public institutions of higher education can better support their students to improve retention and academic success, two key indicators of institutional success.

## Methods

Our research team conducted a baseline survey with a representative sample of 2,282 CUNY students in April 2020 that captured student sociodemographic characteristics and their experiences during the first wave of the COVID-19 pandemic.^8^ Of the 1,035 students who responded to the baseline survey in April 2020 and agreed to be recontacted, we approached a sub-sample of those who (at the time of the baseline survey) lived in any of the 33 neighborhoods defined as most adversely affected by the pandemic, determined by the New York City Mayor’s office.^9^ We reviewed baseline data to group respondents into two categories: 1) “high impact”: those who had any indicators of being highly affected by COVID-19, as defined by measures of mental health (anxiety, depression), food insecurity, housing insecurity, and expected delays to graduation or 2) “low impact”: those who did not demonstrate any indication of mental health problems, food or housing insecurity, nor expected delays to graduation. We reviewed the race/ethnicity, status as a student, NYC borough of residence, gender, and age of respondents to purposively select a diverse group of students in the “high” and “low” impact groups to approach for in-depth interviews (IDIs). To ensure that we captured the perspectives of students who took a leave of absence or had to drop out, we used data from the follow-up survey (fielded to 1,035 students who completed the baseline survey in April 2021; 51% responded) to identify students living in any NYC neighborhood who reported taking a leave or dropping out of CUNY (n=7). We approached a total of 130 students for participation and 38 were enrolled (response rate: 29%). We approached more participants in the “high” (versus “low”) impact group to understand how various risk factors may have affected their ability to persist. We continued enrollment until thematic saturation was reached.

Interviews were conducted virtually via audio or video conferencing from May 17, 2021 to August 20, 2021. During our interviews, the number of COVID-19 cases was declining as the vaccine became widely available. (The delta variant had not yet become widespread in the U.S.) On average, interviews lasted 44 minutes (range: 25-63). The guide contained questions related to academic performance and persistence, physical and mental health, experience with racism/inequities, financial security, and school resources and recommendations; it was iteratively updated as interviews were conducted, and the team discussed these changes. Participants received a $50 cash transfer for their time. In the email confirming payment, students were also directed to our guide to surviving and thriving at CUNY during the pandemic.^10^ The study was approved by the CUNY SPH’s institutional review board (protocol number 695980) and all participants provided informed consent prior to the IDI. Filler words (e.g., um, like) have been removed from the quotes presented below.

### Analysis

Immediately following each IDI, the interviewer recorded field notes documenting: 1) salient interviewee comments that took place post-recording, 2) emerging ideas based on the interview, 3) methodological notes of potential edits to the guide, and 4) reflections including how their feelings about the interviewee and positionality may have affected the interview. These notes were discussed as a team and used to iteratively edit the semi-structured interview guide and probe for emerging ideas in subsequent interviews.

IDIs were transcribed for analysis using Otter transcription software, and then reviewed for completeness by the research team. Three authors who conducted interviews were also analysts (MM, AL, JK). We approached the analysis using methods informed by grounded theory.^11,12^ Four analysts (MM, AL, JL, AK) engaged in qualitatively coding repeating ideas. ^13^ Initially the analytic team coded the same two transcripts to refine the code structure and ensure codes were applied consistently. The analysts then separately coded the remaining transcripts. Throughout the coding and analytic process, questions were discussed and resolved in team meetings. After coding two-thirds of the interviews, analysts revisited all coded transcripts and applied the final code structure to these and the remaining third of interviews. The code application for every transcript was reviewed by two research team members and updates discussed. Throughout the process, each analyst developed memos about emerging analytic ideas. The analytic team engaged in an interactive process of reviewing codes and studying memos to develop themes.^11,13^ Analytic discussions as a team helped to refine the thematic structure. We engaged in a constant comparison across “high” and “low” impact groups to assess similarities and differences in factors that could mitigate the impact of the pandemic on students’ academic engagement and achievement. We also compared students who dropped out or took a leave to those who persisted to assess differences in factors that facilitated or impeded academic engagement. Dedoose software (v. 8.3.43) was used for all qualitative analyses.

## Results

From the baseline survey we found that, of the 38 students included in this sample, 23 (61%) identified as female and 15 (39%) as male (Table 1). There was representation across racial groups with 12 (32%) identifying as Black, 9 (24%) as White, 8 (21%) as Hispanic, and 9 (24%) as another group. From the interview data, we found that almost a third (n=12) were caretakers for younger family members, their own children, or their parents/older relatives. Eleven (29%) had graduated by Fall 2020, four (11%) had graduated and were enrolled in another degree, 16 (42%) were currently enrolled, and 7 (18%) had dropped out or taken a leave of absence. In the entire sample, participants resided in 27 of the 33 impacted neighborhoods; of the seven who dropped out or took a leave of absence three came from less impacted neighborhoods.

**Table 1.** Sample Sociodemographic and Academic Characteristics (n=38)

| Characteristic | N (%) |
| --- | --- |
| Sex |  |
| Female | 23 (61) |
| Male | 15 (39) |
| Race/ethnicity |  |
| Black | 12 (32) |
| White | 9 (24) |
| Hispanic | 8 (21) |
| Another racial/ethnic group | 9 (24) |
| Caretaker |  |
| None | 26 (68) |
| Child(ren) | 4 (11) |
| Family member (siblings, parents) | 8 (21) |
| Academic status |  |
| Graduated | 11 (29) |
| Graduated one program and enrolled in another degree | 4 (11) |
| Currently enrolled | 16 (42) |
| Dropped out/took a leave of absence | 7 (18) |

The pandemic universally altered the course of students’ routines, dramatically shifting the structure of their daily lives. For each student, a constellation of personal, social, and school-related factors created barriers or facilitators to their academic persistence and achievement (Table 2). Their financial stability, social support, mental and physical health, and experience with school and online learning were all connected in ways that affected their capacity to effectively engage in school during the pandemic.

**Table 2.** Facilitators and Barriers to Academic Persistence & Achievement.

|  |  |  |
| --- | --- | --- |
|  | Meeting Essential Needs | Meeting Academic Needs |
| **Facilitators** | Financial stability (e.g., stable income, housing, unemployment benefits)<br>Social safety-net (e.g., family, partner)<br>Professional mental health support (external to school) | Effective online pedagogy<br>-accommodating & accessible professors<br>-synchronous offerings, available class recordings |
|  | ● College student emergency fund | ● College resources (e.g., laptop loaner program, credit/no credit option, career counseling) |
| **Barriers** | ● Disruptive home environment (e.g.- presence of children, family members)<br>● Employment (e.g., needing to work more hours, loss of employment)<br>● Caretaking (e.g., of children, parents)<br>● History of mental health problems (exacerbated during pandemic)<br>● Illness (including COVID)<br>● Disabilities that affect concentration/learning online | ● Online learning<br>-lower quality (vs. in-person courses)<br>-inflexible and/or non-responsive professors<br>● Barriers to accessing to technology<br>● Inaccessible or unsupportive programs/resources<br>● Lacking sense of community at school |

Financial and social support systems were critical to addressing essential needs and allowing students to persist. These supports came from the school, government, or through their personal and social networks. For students whose essential needs were met, the role of faculty and their facilitation of online learning played a critical role in their academic success. In extreme cases where students were trying to manage unmet essential needs, compounded by encounters with faculty perceived to be unsupportive and barriers to online learning, their ability to continue their academic pursuit was considerably undermined.

Depending on their combination of these personal, social, and school-related factors, students fell along a spectrum of academic impact – from dropping out or taking a leave, accepting lower grades given their more limited ability to engage in coursework, maintaining the same level of academic engagement as pre-pandemic, or thriving and performing even better than before.

Heightened anxiety and depression were pervasive among students in this sample. This was often situational, related to fears of themselves or loved ones contracting COVID-19, feeling isolated from their social support system, or being worried about loss of income during the pandemic. Although situational mental health problems alone may not have interfered with students’ ability to remain in school, it likely affected their ability to stay focused and thrive academically.

There were no prominent differences in the manifestation of how personal, social, and school-related factors affected academic achievement by ‘impact’ group. For some, their lived experiences, as told a year later, did not align with what had been their indicators of “high impact” from the baseline survey.

### The Primacy of Meeting Essential Needs

To remain in school, the interviews suggested, students must be able to meet their essential needs for stable housing, food security, physical health, and managing pre-existing mental health problems.

Students with disruptive home environments, loss of employment, or learning disabilities faced barriers to engaging in school. Students with prior mental health problems that were then exacerbated during the pandemic encountered increased challenges to their academic persistence and achievement.

Despite barriers, many were able to address these needs and persevered to continue their schooling. Students with financial and social support to meet their essential needs, were better able to weather the effects of the pandemic (Figure 1). Those who lacked such financial and social support systems were often unable to continue or maintain the same level of engagement in school. One student reflects on her decision to leave school, after she had to work full-time to support her mother and sister:

**Figure 1.**
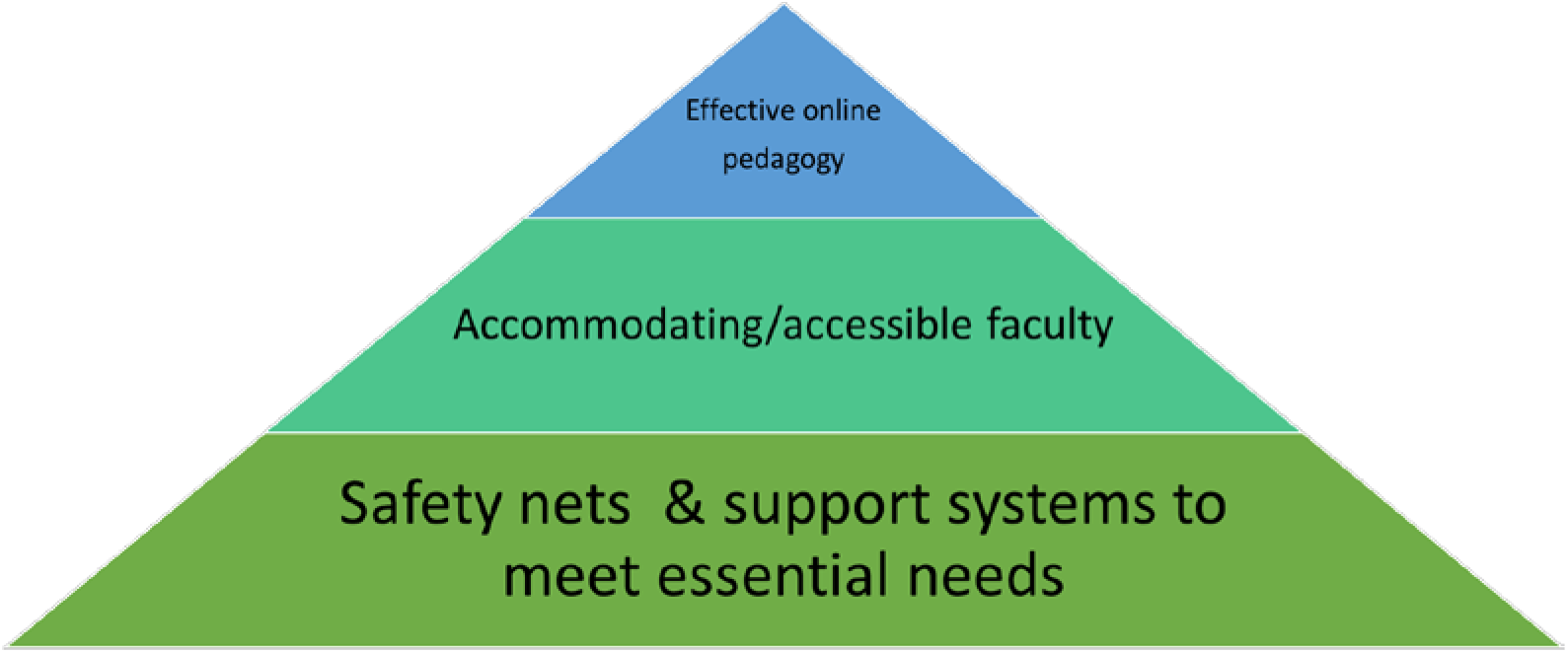
Adapted Maslow’s Pyramid: Hierarchy of Needs for Academic Persistence.

> “I had to decide not to come back to school because I had to care for my family. And I had to take on a full-time job and that mentally would tire me so much. So, won’t be able to attend school and attended where I know I can do good. I didn’t want to go to school, and I knew I’m not doing my best. [Before the pandemic] I was working part-time and then my health got worse. And I had to have a surgery. But after that I was able to bounce back and come back to school and it was great. And then the pandemic hit. And I was like, oh, well, I can’t do that. I have to get a full-time job.”

Many students in our sample lost their employment during the pandemic. In one prominent scenario, this led to a loss of structure, loss of housing, exacerbated mental health problems, or promoted a sedentary lifestyle and weight gain, thus reducing their capacity for schoolwork. In some cases, students’ parents were the ones laid off (as in the situation described above), necessitating the student to work more to help support their family. In another scenario, for those with financial and social safety-nets, the loss of employment was less significant and sometimes came with positive effects such as having more time for schoolwork and more time to plan meals. To demonstrate the disparate paths students faced, we present two scenarios of students who lost their job and main source of income during the pandemic:

*Student #1*: One participant lost a part-time internship that was set to transition to full-time employment in spring 2020. However, she was able to finish her coursework and secure housing and interim income. She described the role of her social support network, particularly her partner, in her ability to stay afloat during this time, noting:

> “I feel really like, blessed to have, my fiancé and how close we were. I lost my job, I had to move out of my apartment, I couldn’t afford to pay for it anymore. And he let me move in with him and his two roommates.”

She credits this, in combination with collecting unemployment and her ability to scale-up alternate sources of income like teaching virtual fitness classes, for her sustained financial stability. She also highlighted the importance of her personal connections, such as family and friends, in mitigating any negative impact on her mental health.

*Student #2*: In contrast, another participant that faced loss of employment at the start of the pandemic reported that this financial strain ultimately led him to drop out of his academic program. The participant described how losing his service industry job created financial challenges that directly affected his ability to stay in school, highlighting the lack of adequate technology to complete his schoolwork, and also increased his child care responsibilities. He explained:

> “I lost a lot financially, and I wasn’t able to get my laptop fixed. I wasn’t able to pay someone to come in my home and watch my son. That was a really huge barrier….If I had a laptop, or I had the money to fix, I definitely would have did school.”

This participant reported being denied unemployment benefits and although he received some financial support from his parents and the school’s established emergency relief grant program, this income was directed to meeting essential household needs. Pandemic-related challenges also affected his sources of social support, noting that:

> “[the pandemic] challenged my relationship. So, me and the mother of my child, we realized we weren’t able to be together. So, you know, we separated. It was just a lot…I was a little depressed at certain times, I did overthink. It definitely was a huge stress.”

Students’ essential needs were often met by resources from their college (e.g., student emergency fund, option to not receive credit/grade, mental health counseling, laptop loaner program) and from public programs (e.g., unemployment benefits, mental health counseling, SNAP benefits). Students who were not able to access needed resources were often forced to drop out, take a leave of absence, or reduce their engagement in school and receive lower grades.

Many students discussed their reasons for not using school-based resources available to meet their based needs. One prominent reason was a lack of awareness that certain resources, such as mental health counseling, were available to them. For students who were aware, they sometimes encountered logistical barriers (e.g., onerous application process, missing deadlines, being passed on to different staff members without resolution). In some cases, their negative pre-pandemic encounters using resources, or interacting with the university made students reluctant to consider using resources during the pandemic. For example, one student reported she had received inadequate advising pre-pandemic and was then distrustful that CUNY mental health counselors could provide her with the necessary support during the pandemic:

> “I felt like anytime I did go to go for academic counseling…there are no appointments…I never really got the greatest of advice, just in regards to certain classes that I was taking. Obviously, no one can tell you what you should do in terms of your academics and your schooling. But I just felt there were certain times that I just wanted pretty general questions answered, and I felt I didn’t get that. So that was frustrating. So during COVID obviously, I wasn’t really eager to reach out to anyone in terms of [school’s name] faculty for help, just because in the past I did not have the greatest of experiences.”

Finally, the perceived lack of sense of community at their college prevented students from reaching out to sources of support within the school; they felt more connected to external resources (such as mental health counseling) and took advantage of their access to those independent resources. The same student reflected on this:

> “…I never felt like I was a part of a community, quite frankly. Everything was either a text or an email. And I’m not saying obviously that those are not great resources, but I just felt like everything that was going on, it was more about the statistics of what was going on, rather than how can we support each other during this time….And that final week of classes…there was really no one to go to, in terms of, ‘Okay, so what’s going to be happening with my classes,’ …I felt that as an individual, there really wasn’t any reassurance…. in terms of overall, [from] the college itself and faculty, I did not have any feeling like you’re part of a community. I did not feel that way at all.”

### Faculty and Online Learning Mattered Next

For students whose essential needs were able to be met by their financial and social support systems (within and outside of the school), and thus able to persist, the role of faculty and their facilitation of online learning was critical to their academic engagement and success. Instructors who were accommodating to special circumstances (e.g., allowing for more time for an assignment) and available (i.e., responsive to emails and questions) were central in facilitating students’ online learning. In contrast, faculty who were inflexible and non-responsive, with what students described as poor online pedagogy, created barriers to academic achievement. To demonstrate these contrasting trajectories, we again present the scenarios of two students who had unsupportive or supportive faculty encounters, respectively.

*Student #3*: One student’s grandparent died from the COVID-19 virus a few months prior to the interview. While she was attending the services, managing her grief, and dealing with a physical ailment, she encountered an unsupportive faculty member. The instructor required proof of the family member’s passing and even with that did not allow the student to make up missed work, affecting her grade. She expressed her frustration trying to get approval to have the late assignments be counted towards her grade for the course:

> “I reached out to the Dean of Students, I reached out to [instructor]. I reached out to numerous people, and I still don’t have permission to send these assignments in. And the fact that I already completed the assignment and sent it, and [instructor] says that ‘Oh, no, the time to do this is over, it passed.’”

The student, who identifies as Black, felt this treatment was racially charged (the faculty member of a different, unspecified race). At the time of the interview, she was taking a heavy course load and considering taking a leave of absence.

*Student #4*: Another student’s father died of COVID-19 several months prior to the interview. Her tuition was fully funded through a scholarship, and she lived with her family, who provided financial support.

When she got infected with COVID-19 and lost her father, instructors offered her flexibility in deadlines:

> “…they [instructors] were very supportive. I know, they said that, of course, I needed to get all the finals in, and all the final exams, or they couldn’t provide me with the grade. And they also offered me, …an incomplete and (to) finish the course next semester, but I didn’t personally want to do that. So I just pushed through, even if it wasn’t always the highest quality work, they still didn’t give me Fs or anything like that…I think what helped me stay on track was just the fact that I was so close to being done.”

For students who were near completion of their degree and had the privilege of support systems, including established peer support pre-pandemic, the imminence of graduating served as a motivating factor.

To address some of the barriers they faced, students offered recommendations for actions their college could take to facilitate their academic success. Increased financial support (for tuition, textbooks, etc.) was prominent in their discussions. Other recommendations included improved communication about existing resources, expanded mental health counseling, fostering a sense of community, and promoting increased flexibility from faculty during the pandemic. Below we integrate students’ recommendations into our recommendations based on the findings.

## Discussion

Our study found that meeting the essential needs of students was central in allowing them to succeed academically. This insight is not new but during the pandemic, meeting these needs became even more vital.^1^ Once such needs are met, the role of faculty was critical in facilitating students’ academic achievement. Thus, public institutions of higher education would benefit from shifting from a narrower focus on academic-related support for individual students to a broader approach addressing students’ essential needs, such as food, housing, financial security, and child care. Such needs should be considered social determinants of academic success. Additionally, institutions of higher education can implement academic policies and procedures to bolster faculty’s ability to facilitate students’ retention and success.

### Recommendations

Given our findings, we propose the following recommendations for public universities to consider during and after the pandemic.

To meet students’ essential needs, institutions can:

- Create or expand student emergency funds and other financial supports (e.g., coverage for textbooks, transportation, technology resources)
- Create a centralized, integrated system of services
- Increase access to long-term, high-quality mental health counseling
- Foster a sense of community within schools, tailored to also meet the needs of new students and students in remote courses
- Allow students to use otherwise unused campus space (to engage in online courses, study, conduct virtual meetings with instructors and peers; to accommodate students who have disruptive home environments)
- Dissolve silos between faculty and student services, to provide an opportunity for faculty to become more aware of resources and connect students to them (given that faculty were students main point of contact)

To meet students’ academic needs, institutions can:

- Create a culture where faculty are expected to be accommodating and flexible (e.g., allowing for extensions when needed, not requiring proof of adverse circumstances, recording classes for students who cannot attend) while upholding the academic integrity of the course material
- Invest in and institutionalize training to improve faculty’s online pedagogy
- Offer options for course mode (online, hybrid, in-person), so students can choose what best suits their situation
- Offer student-centered academic advising to help resolve barriers to learning

The study findings should be interpreted within its limitations. At the time of the interviews, the COVID-19 vaccine was widely available, and the delta variant was not prominent in the U.S. Thus, students’ perspective on the pandemic and its effects may have changed as the fall semester approached and school policies were changing to accommodate the increase in COVID-19 cases. Additionally, students were asked to recall experiences from a year prior, at the height of the pandemic. Their current situation may have influenced how they remembered these early pandemic experiences. While we expected that students from the “low” impact group might identify factors that mitigated the impact of the pandemic, our qualitative analysis did not find any noteworthy differences in barriers and facilitators across these groups. A possible explanation for this finding is that by the end of the first year of the pandemic, most students had developed strategies to find needed resources.

By helping to address students’ essential needs and creating a culture of supportive faculty, institutions of higher education can improve the lives of students and increase academic persistence, achievement, and degree completion.^14^ By approaching policy and intervention development from a perspective that considers students’ holistic lives, during and post-pandemic, leaders of public universities serving low-income students can better serve students, the institution, and the workforce students enter after graduation.

## Data Availability

Data are not yet publicly available.

## Acknowledgements

The authors would like to thank Tara Twiste and Natalya Petroff from the CUNY Office of Applied Research, Evaluation and Data Analysis for their collaboration in aiding with the sampling. We would also like to thank Melissa Carreno and Nida Joseph for transcription and Patricia Lamberson, who helped in her role as Deputy Director of Healthy CUNY.

